# Estimating eligibility for GLP-1 receptor agonists for chronic weight management and cardiovascular disease in Australia

**DOI:** 10.64898/2026.03.17.26348659

**Authors:** Jasmin Castrillon, Chris Schilling, Sharmala Thuraisingam, Michael W Hii, Priya Sumithran, Peter F Choong, Michelle M Dowsey, Cade Shadbolt

## Abstract

**Objectives:** To estimate population-level eligibility for glucagon-like peptide-1 receptor agonist (GLP-1RA) medications among adults in Australia, according to Therapeutic Goods Administration (TGA) approved indications for chronic weight management and secondary prevention of cardiovascular disease.

**Design:** Cross-sectional analysis of data from the Australian Bureau of Statistics 2022 National Health Survey.

**Setting, Participants:** Non-pregnant adults aged ≥18 years who are usual residents of Australia and living in a private dwelling.

**Main outcome measures:** Total number of adults who are eligible for GLP-1RA medications according to TGA approved indications for chronic weight management and secondary prevention of cardiovascular disease, across subgroups defined by body mass index, weight-related comorbidities, and/or socio-demographic factors.

**Results:** In total, 39.7% (95% CI 38.4 – 41.0%) of adults in Australia are eligible for GLP-1RA use for chronic weight management, accounting for 7.8 million (95% CI, 7.6 – 8.1 million) individuals. Among those eligible under this indication, 2.9 million (95% CI 2.7 – 3.1 million) adults had no weight-related comorbidities, 3.3 million (95% CI 3.1 – 3.4 million) adults had at least 1 weight-related comorbidity, and 1.7 million (95% CI 1.6 – 1.8 million) adults had at least 2 weight-related comorbidities. The proportion of adults eligible under this indication varied across clinical and sociodemographic factors. Among those eligible under the chronic weight management indication, up to 338.9 thousand (95% CI 271.3 – 406.5 thousand) adults also meet the indication for secondary prevention of cardiovascular disease.

**Conclusions:** More than one third of Australian adults are eligible to access GLP-1RAs for chronic weight management, with 3.7-4.3% of adults also qualifying according to the indication for established cardiovascular disease. This study provides a valuable reference to allow policy makers to understand the number of adults in Australia that may benefit from access to GLP-1RA medications under a range of coverage scenarios.

**Plain Language Summary:** *The known:* Glucagon-like peptide-1 receptor agonists (GLP-1RA) medications have established benefits for both management of overweight/obesity and the secondary prevention of cardiovascular disease.

*The new:* This study provides the first estimates of the number of adults in Australia who are eligible for treatment with GLP-1RAs across TGA approved indications for chronic weight management and secondary prevention of cardiovascular disease, alongside estimates of eligibility under coverage scenarios based on body mass index and comorbidities.

*The implications:* Our findings offer a policy-relevant reference of the number of adults who could benefit from treatment, supporting advocacy, coverage decisions, and healthcare system planning.

## Introduction

Glucagon-like peptide-1 receptor agonist (GLP-1RA) medications have received regulatory approval by the Therapeutic Goods Administration (TGA) in Australia for indications including type 2 diabetes, chronic weight management, and the secondary prevention of cardiovascular disease [1-4]. The Pharmaceutical Benefits Scheme (PBS) subsidises semaglutide and dulaglutide for individuals with type 2 diabetes that has not adequately responded to other antihyperglycemic medications including sodium-glucose cotransporter 2 inhibitors [5, 6]. No GLP-1RA medications are currently subsidised by the PBS for obesity or secondary prevention of cardiovascular disease. However, based on demonstration of cardiovascular benefits [7], the Pharmaceutical Benefits Advisory Committee recently recommended subsidising semaglutide for adults with established cardiovascular disease and obesity, [8] indicating that subsidies may be limited to individuals with a body mass index (BMI) of 35kg/m^2^ or higher due to the cost of the medication [8].

Despite ongoing national and international debate about appropriate coverage of GLP-1RA medications for indications beyond diabetes [9, 10], the total number of adults in Australia who are currently eligible to use these medications for chronic weight management or secondary prevention of cardiovascular disease remains unclear. In a recent international study, 27% of adults across 99 countries (not including Australia) were eligible to use GLP-1RA medications according to their approved indication for obesity [11]. Notably, these estimates varied widely even between high-income countries, further highlighting the need for reliable local estimates. In this study, we aim to provide nationally representative estimates of the total number of adults currently eligible according to TGA approved indications for GLP-1RAs for chronic weight management and secondary prevention of established cardiovascular disease. We also aim to provide estimates of eligibility for these medications across sub-populations defined by key sociodemographic and clinical characteristics and estimate the number of adults that may be eligible for access to these medications under a range of coverage scenarios.

## Methods

### Data source and sample selection

Our sample included all non-pregnant adults aged 18 years or older included in the 2022 National Health Survey conducted by the Australian Bureau of Statistics [12]. Pregnant adults were excluded due to both the absence of accurate BMI measurements and pregnancy being an explicit contraindication to GLP-1RA use. The National Health Survey collects data from a sample of randomly selected households across Australia [13]. Survey respondents are interviewed to collect information about sociodemographics, health conditions, and physical measurements [13]. The Australian Bureau of Statistics provides calibrated person-level survey weights to enable unbiased nationally representative estimates of its target population of all usual residents in Australia living in private dwellings, which account for both the sampling design and non-response (43.3%) [13].

### Identification of eligible individuals Chronic weight management

Individuals that are eligible for GLP-1RA medications for chronic weight management were identified based on the approved indications outlined by the TGA. Currently, semaglutide and tirzepatide are the only available medications with regulatory approval for this indication [1, 3]. We defined eligible individuals as adults with a BMI ≥30 kg/m^2^ or a BMI ≥27 kg/m^2^ to <30 kg/m^2^ and at least one weight-related comorbidity including, hypertension, dyslipidaemia, cardiovascular disease, or type 2 diabetes. As weight-related comorbidities were not clearly defined in the TGA approval, we limited these conditions to those used to determine eligibility in key Phase 3 trials of these medications [14-17]. Obstructive sleep apnoea was not included in our analysis, as the National Health Survey does not collect information on this condition. Consistent with the eligibility criteria of these key trials, we employed the broad criteria developed by the National Health Survey (e.g. angina, myocardial infarction, ischaemic heart diseases, stroke, other cerebrovascular diseases, oedema, heart failure, and other vascular disorders) to define cardiovascular disease.

### Secondary prevention of cardiovascular disease

Currently, semaglutide is the only GLP-1RA medication approved in Australia for secondary prevention of cardiovascular disease in adults with overweight or obesity [1]. We defined individuals as eligible according to this indication if they were adults with established cardiovascular disease and a BMI ≥27 kg/m^2^, without diabetes. Established cardiovascular disease was defined in the clinical trial used to support this indication as having had a prior myocardial infarction, prior stroke, or symptomatic peripheral arterial disease [7]. Our primary definition of *confirmed established cardiovascular disease* included only individuals with a prior stroke and/or myocardial infarction, which were conditions specifically recorded by the survey. However, the National Health Survey does not provide specific information on diagnoses of symptomatic peripheral arterial disease, which is grouped within the broader category of diseases of arteries, arterioles, and capillaries (ICD-10-CM Diagnosis Code I70-I79). To examine the impact of our more conservative primary definition, we classified individuals as having *possible established cardiovascular disease* if they reported a condition coded to the broader category of diseases of arteries, arterioles, and capillaries without meeting our primary definition of established cardiovascular disease (i.e. no prior myocardial infarction or stroke).

### Statistical analysis

We estimated the total number of non-pregnant adults in Australia who are eligible and not eligible for GLP-1RAs for chronic weight management across several sociodemographic and clinical characteristics including age, sex, education (secondary or below, certificate/diploma, university-level), weight-related comorbidities, and household income quintiles. We then estimated the proportion of individuals within each of these sub-populations that were eligible for GLP-1RAs for chronic weight management. To examine population-level eligibility under a range of different coverage scenarios, we calculated the total number of adults in Australia eligible for GLP-1RAs for chronic weight management across subgroups defined by both BMI categories and number of weight-related comorbidities. Among those eligible for chronic weight management, we then estimated the total number of adults eligible and not eligible for GLP-1RAs for secondary prevention of established cardiovascular disease across subgroups defined by BMI categories.

All analyses used person-level survey weights to derive nationally representative estimates, with jackknife replicate weights used to estimate corresponding 95% confidence intervals [13, 18]. All data was analysed in the ABS DataLab using Stata V.18.0 (StataCorp).

### Ethics statement

This study was approved by The University of Melbourne Human Research Ethic Committee (Reference number: 2025-32594-66417-3).

## Results

In total, 13,039 individuals were included in the unweighted sample, representing 19.7 million (95% CI 19.7 – 19.7 million) non-pregnant adults (Table). After weighting, an estimated 39.7% (95% CI 38.4 – 41.0%) of adults were eligible for GLP-1RA use for chronic weight management, accounting for 7.8 million (95% CI, 7.6 – 8.1 million) adults. The proportion of adults eligible for GLP-1RA use for chronic weight management varied across clinical and sociodemographic factors. Of note, a greater proportion of adults was eligible among those with the lowest household income (44.4%; 95% CI 41.6 – 47.3%) when compared to those with the highest household income (36.7%; 95% CI 33.9 – 39.6%). Around two-thirds of individuals across all assessed weight-related comorbidities were eligible for GLP-1RAs under the chronic weight management indication, with eligible adults accounting for 70.3% (95% CI, 66.1 - 74.1%) of those with type 2 diabetes, 65.6% (95% CI, 63.5 – 67.6%) of those with hypertension, and 64.8% (95% CI, 60.6 – 68.8%) of those with dyslipidaemia.

Of the 7.8 million adults eligible for GLP-1RA use for chronic weight management, 2.9 million (95% CI 2.7 – 3.1 million) adults had no weight-related comorbidities, 3.3 million (95% CI 3.1 – 3.4 million) adults had at least 1 weight-related comorbidity, and 1.7 million (95% CI 1.6 – 1.8 million) adults had at least 2 weight-related comorbidities (Figure 1; Supporting Information, Table 1). Additionally, among these eligible individuals, 1.6 million (95% CI 1.4 – 1.7 million) had a BMI between 27-29.9kg/m^2^, 3.8 million (95% CI 3.6 – 3.9 million) had a BMI between 30-34.9kg/m^2^, 1.6 million (95% CI 1.5 – 1.7 million) had a BMI between 35-39.9kg/m^2^, and 909.0 thousand (95% CI 805.8 - 1012.1 thousand) had a BMI of 40kg/m^2^ or higher. Figure 1 presents the total number of eligible adults for each combination of BMI category and number of weight-related comorbidities.

**Figure 1.**
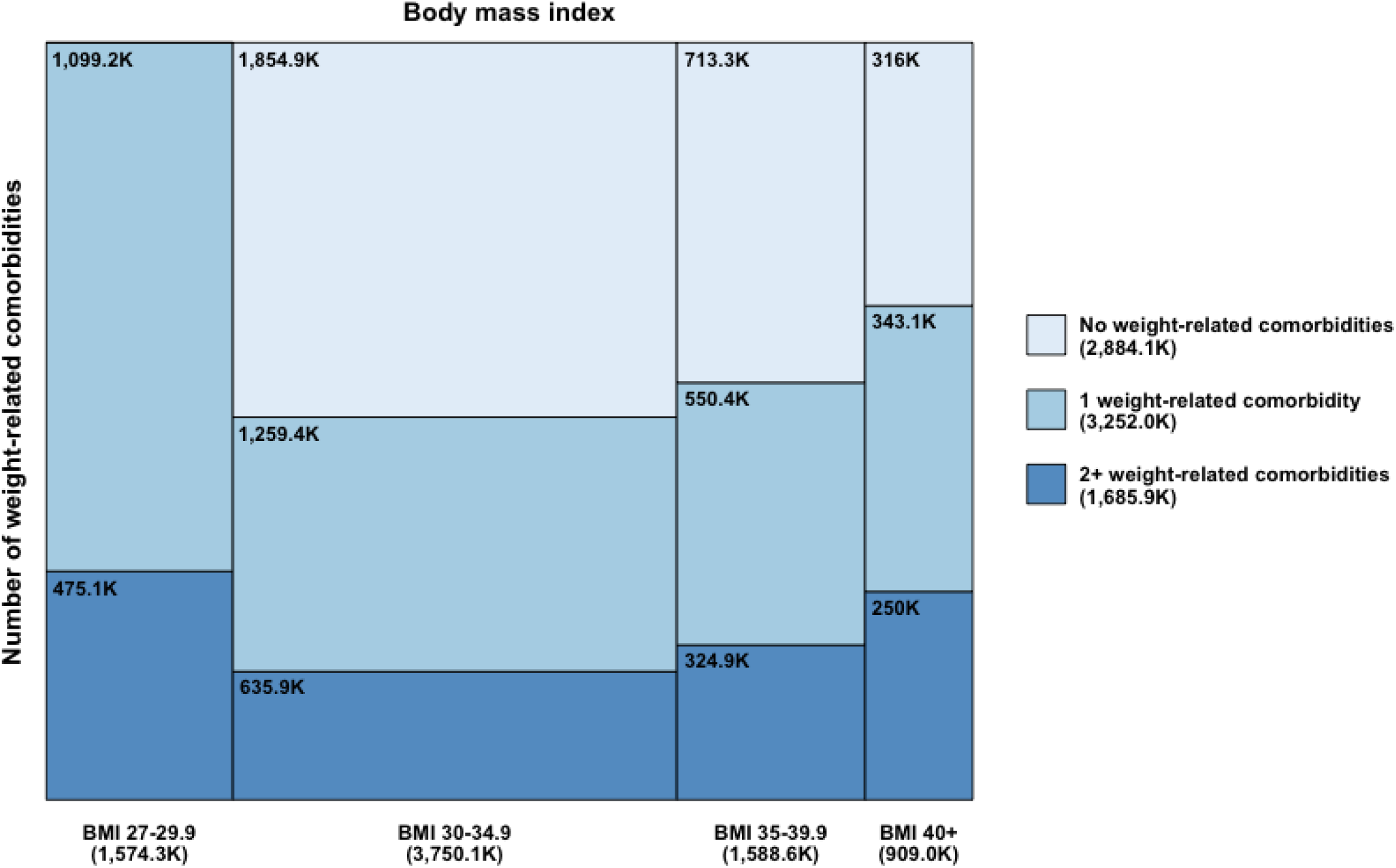
Estimated number of adults eligible for GLP-1RA use for chronic weight management by body mass index and number of weight-related comorbidities Abbreviations: BMI, body mass index. The size of each tile represents the proportion of adults within each BMI category, with or without weight-related comorbidities. The eligible sample includes non-pregnant adults with BMI ≥ 30kg/m2 or BMI ≥27kg/m2 to <30kg/m2 and at least one weight-related comorbidity. Excludes adults with type 1 diabetes.

**Table 1.** Characteristics of Australian adults eligible for glucagon-like peptide-1 receptor agonist use for chronic weight management.

|  | Estimated population, No. (95% CI) (Thousands) |  |  |  |
| --- | --- | --- | --- | --- |
| Characteristics | Overall<sup>d</sup> | Eligible<sup>e</sup> | Non-eligible | Proportion eligible (95% CI) |
| Unweighted, n | 13039 | 5452 | 7587 |  |
| Weighted, n (Million) | 19.7 (19.7 to 19.7) | 7.8 (7.6 to 8.1) | 11.9 (11.6 to 12.1) | 39.7 (38.4 to 41.0) |
| **Age** |  |  |  |  |
| 18-29 | 3813.3 (3696.0 to 3930.6) | 841.9 (726.6 to 957.2) | 2971.4 (2849.3 to 3093.5) | 22.1 (19.4 to 25.0) |
| 30-44 | 5496.4 (5381.1 to 5611.8) | 1916.6 (1784.3 to 2048.9) | 3579.8 (3425.6 to 3734.0) | 34.9 (32.6 to 37.3) |
| 45-64 | 6197.1 (6192.0 to 6202.2) | 2874.7 (2735.7 to 3013.7) | 3322.3 (3181.6 to 3463.1) | 46.4 (44.1 to 48.7) |
| 65+ | 4196.4 (4196.4 to 4196.4) | 2188.8 (2113.5 to 2264.0) | 2007.7 (1932.4 to 2082.9) | 52.2 (50.4 to 53.9) |
| **Sex** |  |  |  |  |
| Male | 9714.5 (9714.4 to 9714.6) | 4041.3 (3881.3 to 4201.4) | 5673.2 (5513.1 to 5833.2) | 41.6 (40.0 to 43.3) |
| Female | 9988.7 (9956.0 to 10021.4) | 3780.7 (3624.7 to 3936.7) | 6208.0 (6050.0 to 6366.0) | 37.8 (36.3 to 39.4) |
| **Clinical History** |  |  |  |  |
| T2D | 1176.7 (1063.4 to 1289.9) | 826.8 (741.4 to 912.2) | 349.9 (287.2 to 412.5) | 70.3 (66.1 to 74.1) |
| Hypertension | 6368.8 (6148.3 to 6589.4) | 4175.4 (3990.9 to 4360.0) | 2193.4 (2037.0 to 2349.7) | 65.6 (63.5 to 67.6) |
| Dyslipidaemia | 2216.3 (2055.1 to 2377.6) | 1435.9 (1289.7 to 1582.2) | 780.4 (679.0 to 881.8) | 64.8 (60.6 to 68.8) |
| Previous myocardial infarction | 434.0 (356.0 to 511.9) | 288.1 (224.8 to 351.3) | 145.9 (106.5 to 185.3) | 66.4 (58.6 to 73.3) |
| Previous stroke | 193.2 (156.6 to 229.8) | 113.7 (81.6 to 145.8) | 79.4 (53.1 to 105.8) | 58.9 (46.8 to 70.0) |
| Diseases of arteries, arterioles, and capillaries | 171.2 (129.6 to 212.8) | 98.2 (72.8 to 123.7) | 73.0 (40.3 to 105.7) | 57.4 (44.3 to 69.5) |
| Other CVD<sup>a</sup> | 849.8 (756.2 to 943.4) | 545.4 (470.1 to 620.8) | 304.4 (248.2 to 360.6) | 64.2 (58.7 to 69.3) |
| Osteoarthritis | 2095.7 (1976.0 to 2215.4) | 1222.8 (1130.9 to 1314.7) | 872.9 (790.4 to 955.4) | 58.3 (55.3 to 61.3) |
| Back pain and other dorsopathies<sup>b</sup> | 3932.8 (3741.0 to 4124.7) | 1926.6 (1775.1 to 2078.0) | 2006.3 (1889.5 to 2123.0) | 49.0 (46.6 to 51.4) |
| **Income<sup>c</sup>** |  |  |  |  |
| 1st quintile (lowest) | 2742.4 (2575.7 to 2909.2) | 1218.2 (1116.6 to 1319.9) | 1524.2 (1396.4 to 1651.9) | 44.4 (41.6 to 47.3) |
| 2nd quintile | 3142.7 (3007.8 to 3277.6) | 1453.9 (1356.9 to 1551.0) | 1688.8 (1565.6 to 1811.9) | 46.3 (43.5 to 49.0) |
| 3rd quintile | 3539.0 (3358.8 to 3719.2) | 1342.4 (1227.5 to 1457.2) | 2196.6 (2049.5 to 2343.8) | 37.9 (35.3 to 40.6) |
| 4th quintile | 3994.6 (3738.2 to 4251.0) | 1530.0 (1390.3 to 1669.7) | 2464.6 (2260.8 to 2668.4) | 38.3 (35.5 to 41.1) |
| 5th quintile (highest) | 4223.3 (4046.9 to 4399.8) | 1548.8 (1416.6 to 1681.0) | 2674.5 (2505.4 to 2843.7) | 36.7 (33.9 to 39.6) |

| Education |  |  |  |  |
| --- | --- | --- | --- | --- |
| Secondary education or below | 7604.2 (7493.3 to 7715.0) | 3232.4 (3049.4 to 3415.3) | 4371.8 (4218.3 to 4525.2) | 42.5 (40.4 to 44.7) |
| Certificate/Diploma | 4594.7 (4480.7 to 4708.7) | 2030.4 (1915.8 to 2145.1) | 2564.3 (2443.2 to 2685.3) | 44.2 (42.0 to 46.4) |
| University-level education | 7037.3 (6903.1 to 7171.5) | 2354.9 (2201.4 to 2508.3) | 4682.4 (4530.0 to 4834.7) | 33.5 (31.5 to 35.5) |
| Other | 467.1 (388.2 to 546.1) | 204.3 (159.2 to 249.5) | 262.8 (207.2 to 318.4) | 43.7 (37.2 to 50.5) |

Abbreviations: CVD, cardiovascular disease; NA, not applicable; T2D, type 2 diabetes.
a Includes angina, other ischaemic heart diseases, other cerebrovascular diseases, oedema, and heart failure.
b Includes back pain or problems, sciatica, disc disorders, and curvature of the spine.
c Total current weekly household income from all sources. A total of 1070 adults did not report total current weekly household income.
d Includes non-pregnant adults aged 18 years and older.
e Includes non-pregnant adults aged 18 years and older with a BMI $\geq 30\text{kg/m}^2$ or BMI $\geq 27\text{kg/m}^2$ to $<30\text{kg/m}^2$ and at least one weight-related comorbidity. Excludes adults with type 1 diabetes.

In total, up to 338.9 thousand (95% CI 271.3 – 406.5 thousand) Australian adults are eligible for GLP-1RA medications for established cardiovascular disease. Of these, 285.7 thousand (95% CI, 224.4 – 346.9 thousand) met our definition of confirmed established cardiovascular disease and 53.2 thousand (95% CI, 33.2 – 73.3 thousand) additionally met our definition of possible established cardiovascular disease (Figure 2; Supporting Information, Table 2). In total, 7.5 million (95% CI, 7.2 – 7.7 million) adults eligible for the chronic weight management are not eligible for secondary prevention of established cardiovascular disease. For those confirmed as eligible under this indication, 92.0 thousand (95% CI 56.1 - 127.8 thousand) adults had a BMI between 27-29.9kg/m^2^, 119.7 thousand (95% CI 76.3 - 163.0 thousand) adults had a BMI between 30-34.9kg/m^2^, 41.7 thousand (95% CI 21.7 - 61.7 thousand) adults had a BMI between 35-39.9kg/m^2^, and 32.3 thousand (95% CI 14.4 to 50.2 thousand) adults had a BMI of 40kg/m^2^ or higher. Figure 2 presents the total number of eligible adults with confirmed and possible established cardiovascular disease across BMI categories.

**Figure 2.**
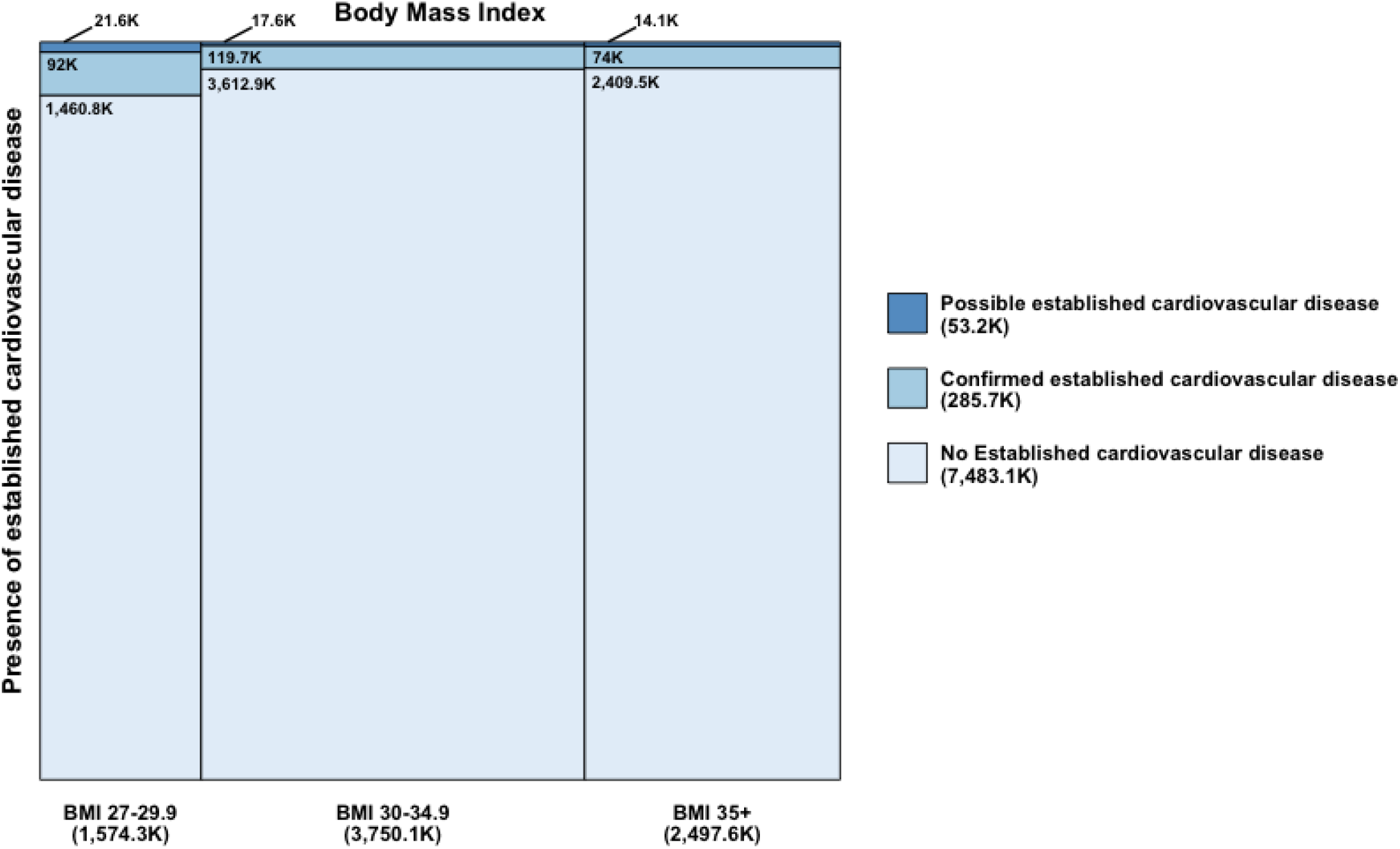
Estimated number of adults eligible for GLP-1RA use for chronic weight management by body mass index and presence of established cardiovascular disease Abbreviations: BMI, body mass index. The size of each tile represents the proportion of adults within each BMI category, with or without confirmed or possible established cardiovascular disease. The eligible sample includes non-pregnant adults with BMI ≥ 30kg/m2 or BMI ≥27kg/m2 to <30kg/m2 and at least one weight-related comorbidity. Excludes adults with type 1 diabetes. Confirmed established cardiovascular disease includes non-pregnant adults with a BMI ≥27 kg/m^2^, prior myocardial infarction or prior stroke, and without diabetes. Possible established cardiovascular disease includes non-pregnant adults with a BMI ≥27 kg/m^2^, prior myocardial infarction, prior stroke, or diseases of arteries, arterioles, or capillaries, and without diabetes.

## Discussion

This study provides the first published estimates of population-level eligibility for GLP-1RA medications in Australia, indicating that 39.7% of adults are eligible under the TGA indication for chronic weight management. Of those eligible, up to 4.3% also meet the criteria for secondary prevention of established cardiovascular disease. These figures are slightly lower than has been reported in previous studies examining other high-income countries, where approximately 41.8% of adults are estimated to be eligible to use this class of medications for weight management [11]. In addition, the nationwide estimates of the number of adults meeting these approved indications under various coverage scenarios provides useful information to healthcare professionals regarding how many patients may benefit from treatment, as well as to policymakers involved in determining criteria for potential subsidies for these medications.

Overall, our findings indicate that more than one third of Australian adults are eligible for GLP-1RA use for chronic weight management. Without PBS-subsidy, the cost of the GLP-1RA semaglutide, at the recommended maintenance dose of 2.4mg, is approximately $AUD 5200 per person annually. Estimating total national spending relies on several factors, such as dosing, price negotiations with manufacturers, and subsidy conditions. However, if coverage were extended to all eligible adults who meet the current approved indication for chronic weight management at the current private prescription price, total annual government spending could reach up to $AUD 40.7 billion. Such an estimate is extreme, as it would require complete uptake among all eligible individuals in the absence of lower negotiated prices. As a point of comparison, the total direct and indirect economic impact of overweight and obesity in Australia was estimated at approximately $AUD 38 billion per year in 2019 [19]. Even far lower levels of uptake and successful pricing negotiations would have significant economic implications for federal budgets.

Limiting subsidised access based on an individual’s BMI and the presence of weight-related comorbidities may be one approach to mitigating high costs. Elevated BMI is associated with a higher risk of developing several conditions, including cardiovascular disease and type 2 diabetes [20, 21]. Our findings indicate that limiting coverage to adults with a BMI exceeding 35kg/m^2^ would provide subsidised access to approximately 2.5 million adults. A further reduction could be achieved by limiting eligibility to adults with a BMI >35kg/m^2^ and at least one weight-related comorbidity, resulting in potential subsidised access to an estimated 1.5 million adults. While such approaches may constrain costs, post-hoc analyses of the STEP and SURMOUNT trials have suggested that both semaglutide and tirzepatide are effective in reducing body weight across all BMI subgroups ≥27kg/m^2^ [22, 23]. Moreover, even in the absence of associated chronic diseases, obesity can substantially impair health and function [24].

Importantly, our findings also demonstrate that eligibility is not evenly distributed across socioeconomic groups. Nearly 45% of individuals living in low-income households currently meet criteria for these medications for weight management, with high out-of-pocket costs associated with private prescriptions raising concerns about inequitable access. Prioritising coverage to those at greatest risk of adverse health outcomes, including consideration of social determinants of health, may better facilitate equitable access to these medications. Such an approach would be consistent with recent WHO guidelines which emphasise the urgent need for evidence-based, equity-focused strategies to expand access to these medications for adults with obesity [25].

Furthermore, our study suggests that approximately 338.9 thousand adults would be eligible for semaglutide under its current approval for secondary prevention of cardiovascular disease. Based on recent PBAC recommendations [8], limiting semaglutide eligibility to adults with established cardiovascular disease and a BMI >35kg/m^2^ would reduce the maximum expected number of subsidised individuals to approximately 88.1 thousand adults. While this approach aids in reducing total spending, subgroup analysis of the SELECT trial indicates cardioprotective benefits of semaglutide across all BMI groups greater than 27kg/m^2^, with the largest observed reduction in major adverse cardiovascular events among adults with a BMI of less than 35kg/m^2^ [26]. This emphasises the need for caution when limiting coverage based on BMI, as doing so may result in limiting access for patients who are likely to benefit from treatment based on available data from large, randomised trials [7].

### Limitations

This study has several limitations. First, we did not have access to data on obstructive sleep apnoea diagnoses, which is one of the weight-related comorbidities used to determine eligibility for these medications. Because of this, estimates of the number of people eligible for these medications for weight management with a BMI less than 30kg/m^2^ are likely to be somewhat conservative. Second, the National Health Survey aggregates data on vascular diseases into a broader category of diagnoses related to arteries, arterioles, and capillaries (ICD-10-CM Diagnosis Code I70-I79). Because of this, we could not specifically identify individuals with symptomatic peripheral arterial disease in our analysis of those eligible for secondary prevention of established cardiovascular disease. However, our analysis classifying individuals with this broader category of vascular conditions as potentially meeting this indication provides an estimate of the upper limit of how many adults may be eligible for GLP-1RAs for cardiovascular risk reduction. Third, there are several potential contradictions to GLP-1RA medications that could not be reliably identified with the data available. Fourth, some survey respondents had missing data on physical measurement. For these participants, we relied upon values imputed by the Australian Bureau of Statistics using established processes based upon self-reported height, weight, and other key predictive factors to provide representative estimates of BMI across the entire survey population [13].

## Conclusion

Approximately 7.8 million Australian adults are currently eligible for GLP-1RA medications according to approved indications for chronic weight management, with up to 3.7-4.3% of adults also meeting the approved indication for secondary prevention of established cardiovascular disease. Of note, almost 45% of individuals living in low-income households are currently eligible to use these medications for weight management, though prohibitive costs for private market prescriptions raises concerns about inequities in access to these effective treatments. Acknowledging that broad PBS-subsidised access for all eligible adults would pose challenges due to the high cost of treatment, this study provides valuable insight to allow clinicians and policy makers to understand the number of adults in Australia that may benefit from access to GLP-1RA medications under a range of possible coverage scenarios.

## Data Availability

All aggregated data in the present study are available upon reasonable request to the authors. All detailed microdata are available only to approved researchers through the Australian Bureau of Statistics DataLab and are not publicly available.

## Author Contributions

JC: Conceptualization; Formal analysis; Writing – original draft. CSch, ST, MWH, PS, PFC, MMD and CSha: Supervision; Conceptualization; Writing-review and editing.

## References

1 Australian Government Department of Health, Disability and Ageing Therapeutic Goods Administration, Australian Product Information Wegovy® (semaglutide) solution for injection (TGA, 2024), Available from: https://www.ebs.tga.gov.au/ebs/picmi/picmirepository.nsf/pdf?OpenAgent=&id=CP-2022-PI-01930-1

2 Australian Government Department of Health, Disability and Ageing Therapeutic Goods Administration, Australian Product Information Ozempic® (semaglutide) solution for injection (TGA, 2019), Available from: https://www.ebs.tga.gov.au/ebs/picmi/picmirepository.nsf/pdf?OpenAgent=&id=CP-2019-PI-01881-1

3 Australian Government Department of Health, Disability and Ageing Therapeutic Goods Administration, Australian Product Information MOUNJARO® (TIRZEPATIDE) SOLUTION FOR INJECTION (TGA, 2025), Available from: https://www.ebs.tga.gov.au/ebs/picmi/picmirepository.nsf/pdf?OpenAgent=&id=CP-2023-PI-02114-1

4 Australian Government Department of Health, Disability and Ageing Therapeutic Goods Administration, AUSTRALIAN PRODUCT INFORMATION – TRULICITY (DULAGLUTIDE RCH) AUTOINJECTOR (TGA, 2025), Available from: https://www.ebs.tga.gov.au/ebs/picmi/picmirepository.nsf/pdf?OpenAgent=&id=CP-2015-PI-01412-1

5 The Pharmaceutical Benefits Scheme, Semaglutide, https://www.pbs.gov.au/medicine/item/12075m-12080t

6 The Pharmaceutical Benefits Scheme, Dulaglutide, https://www.pbs.gov.au/medicine/item/11364D

7 A. M. Lincoff, K. Brown-Frandsen, H. M. Colhoun, et al., “Semaglutide and cardiovascular outcomes in obesity without diabetes,” New England Journal of Medicine, no. 24 (2023): 2221–2232.

8 The Pharmaceutical Benefits Scheme, Recommendations made by the PBAC – November 2025, Available from: https://www.pbs.gov.au/industry/listing/elements/pbac-meetings/pbac-outcomes/2025-11/pbac-web-outcomes-11-2025-v2.pdf

9 J. L. Dellgren, G. Persad, and E. J. Emanuel, “International coverage of GLP-1 receptor agonists: a review and ethical analysis of discordant approaches,” The Lancet, no. 10455 (2024): 902–906.

10 E. J. Emanuel, J. L. Dellgren, M. S. McCoy, and G. Persad, “Fair allocation of GLP-1 and dual GLP-1-GIP receptor agonists,” New England Journal of Medicine, no. 20 (2024): 1839–1842

11 S. G. K. Yoo, F. Teufel, M. Theilmann, et al., “GLP-1 receptor agonists for obesity: eligibility across 99 countries,” The Lancet Diabetes & Endocrinology, no. 2 (2026): 105–08.

12 Australian Bureau of Statistics (ABS) (2022) Detailed Microdata: National Health Survey, ABS DataLab. Findings based on use of ABS data.

13 Australian Bureau of Statistics, National Health Survey methodology (ABS, 2023), https://www.abs.gov.au/methodologies/national-health-survey-methodology/2022

14 J. P. Wilding, R. L. Batterham, S. Calanna, et al., “Once-weekly semaglutide in adults with overweight or obesity,” New England Journal of Medicine, no. 11 (2021): 989–1002.

15 M. Davies, L. Færch, O. K. Jeppesen, et al., “Semaglutide 2· 4 mg once a week in adults with overweight or obesity, and type 2 diabetes (STEP 2): a randomised, double-blind, double-dummy, placebo-controlled, phase 3 trial,” The Lancet, no. 10278 (2021): 971–984.

16 A. M. Jastreboff, L. J. Aronne, N. N. Ahmad, et al., “Tirzepatide once weekly for the treatment of obesity,” New England Journal of Medicine, no. 3 (2022): 205–216.

17 W. T. Garvey, J. P. Frias, A. M. Jastreboff, et al., “Tirzepatide once weekly for the treatment of obesity in people with type 2 diabetes (SURMOUNT-2): a double-blind, randomised, multicentre, placebo-controlled, phase 3 trial,” The Lancet, no. 10402 (2023): 613–626.

18 Australian Bureau of Statistics, Microdata and TableBuilder: National Health Survey, https://www.abs.gov.au/statistics/microdata-tablebuilder/available-microdata-tablebuilder/national-health-survey#further-information

19 World Obesity, Australia Economic impact of overweight and obesity (World Obesity, 2023), https://data.worldobesity.org/economic-impact-new/countries/AU.pdf

20 S. S. Khan, H. Ning, J. T. Wilkins, et al., “Association of body mass index with lifetime risk of cardiovascular disease and compression of morbidity,” JAMA cardiology, no. 4 (2018): 280–287.

21 A. Must, J. Spadano, E. H. Coakley, A. E. Field, G. Colditz, and W. H. Dietz, “The Disease Burden Associated With Overweight and Obesity,” JAMA, no. 16 (1999): 1523–1529.

22 B. M. McGowan, A. Houshmand-Oeregaard, P. N. Laursen, N. Zeuthen, and J. Baker-Knight, “Impact of BMI and comorbidities on efficacy of once-weekly semaglutide: Post hoc analyses of the STEP 1 randomized trial,” Obesity, no. 4 (2023): 990–999.

23 C. Le Roux, L. J. Aronne, F. Jaouimaa, et al., “TS12.01 Tirzepatide for the treatment of obesity reduced body weight across body mass index categories: post hoc analysis of the SURMOUNT 1-4 trials,” Obes Facts, (2024): 27.

24 F. Rubino, D. E. Cummings, R. H. Eckel, et al., “Definition and diagnostic criteria of clinical obesity,” The Lancet Diabetes & Endocrinology, no. 3 (2025): 221–262.

25 World Health Organization, WHO guideline on the use of glucagon-like peptide-1 (GLP-1) therapies for the treatment of obesity in adults (WHO, 2025), Available from: <https://files.magicapp.org/guideline/5f0963f2-daa8-4d20-b0fc-eab3bcf69c48/published_guideline_10802-1_0.pdf>

26 J. Deanfield, A. M. Lincoff, S. E. Kahn, et al., “Semaglutide and cardiovascular outcomes by baseline and changes in adiposity measurements: a prespecified analysis of the SELECT trial,” The Lancet, no. 10516 (2025): 2257–2268.

