## Supporting Information for "Estimating eligibility for GLP-1 receptor agonists for chronic weight management and cardiovascular disease in Australia"

### **Table 1.** Cross-tabulation of the estimated number of adults eligible for glucagon-like peptide-1 receptor agonist use for chronic weight management, by body mass index and number of weight-related comorbidities

|  | **BMI category, n (95% CI) (Thousands)** | | | |  |
| --- | --- | --- | --- | --- | --- |
| **No. of weight-related comorbidities, n (95% CI) (Thousands)** | **27-29.9** | **30-34.9** | **35-39.9** | **40+** | **Total** |
| **None** | NA | 1854.9 (1723.7 to 1986.1) | 713.3 (618.2 to 808.4) | 316.0 (247.1 to 384.8) | 2884.1 (2716.8 to 3051.5) |
| **1** | 1099.2 (993.3 to 1205.1) | 1259.4 (1135.7 to 1383.0) | 550.4 (486.0 to 614.7) | 343.1 (279.1 to 407.0) | 3252.0 (3056.8 to 3447.2) |
| **2+** | 475.1 (417.3 to 532.9) | 635.9 (554.6 to 717.1) | 324.9 (266.5 to 383.3) | 250.0 (202.9 to 297.0) | 1685.9 (1567.2 to 1804.5) |
| **Total** | 1574.3 (1446.4 to 1702.3) | 3750.1 (3579.1 to 3921.2) | 1588.6 (1459.2 to 1717.9) | 909.0 (805.8 to 1012.1) | 7822.0 (7571.2 to 8072.8) |

Abbreviations: NA, not applicable. The eligible sample includes non-pregnant adults with BMI ≥ 30kg/m2 or BMI ≥27kg/m2 to <30kg/m2 and at least one weight-related comorbidity. Excludes adults with type 1 diabetes.

### **Table 2.** Cross-tabulation of the estimated number of adults eligible for glucagon-like peptide-1 receptor agonist use for chronic weight management, by body mass index and presence of established cardiovascular disease

|  | **BMI category, n (95% CI) (Thousands)** | | |  |
| --- | --- | --- | --- | --- |
| **Established cardiovascular disease, n (95% CI) (Thousands)** | **27-29.9** | **30-34.9** | **35+** | **Total** |
| None | 1460.8 (1342.6 to 1579.1) | 3612.9 (3435.3 to 3790.5) | 2409.5 (2252.7 to 2566.2) | 7483.1 (7232.5 to 7733.8) |
| Confirmed established cardiovascular disease | 92.0 (56.1 to 127.8) | 119.7 (76.3 to 163.0) | 74.0 (45.4 to 102.6) | 285.7 (224.4 to 346.9) |
| Possible established cardiovascular disease | 21.6 (8.1 to 35.0) | 17.6 (6.2 to 29.0) | 14.1 (5.5 to 22.6) | 53.2 (33.2 to 73.3) |
| **Total** | 1574.3 (1446.4 to 1702.3) | 3750.1 (3579.1 to 3921.2) | 2497.6 (2338.3 to 2656.8) | 7822.0 (7571.2 to 8072.8) |

The eligible sample includes non-pregnant adults with BMI ≥ 30kg/m2 or BMI ≥27kg/m2 to <30kg/m2 and at least one weight-related comorbidity. Excludes adults with type 1 diabetes. Confirmed established cardiovascular disease includes non-pregnant adults with a BMI ≥27 kg/m^2^, prior myocardial infarction or prior stroke, and without diabetes. Possible established cardiovascular disease includes non-pregnant adults with a BMI ≥27 kg/m^2^, prior myocardial infarction, prior stroke, or diseases of arteries, arterioles, or capillaries, and without diabetes.
